# Who uses mental health support forums, and why? Triangulating findings from surveys, interviews, and forum posts

**DOI:** 10.1101/2025.05.11.25327409

**Authors:** Zoe Glossop, Anna Lindroos Čermáková, Paul Marshall, Paul Rayson, Heather Robinson, Elena Semino, Fiona Lobban

## Abstract

Mental health services in the UK are increasingly pressured with long waiting times. Meanwhile, online forums for mental health, set up by charities, NHS services, and individual volunteers, have increased in popularity. Little is known, however, about who is using them and why. This study aimed to investigate this, using multiple methods to capture different types of users.

A mixed-methods approach was used, with participants recruited from seven UK-based mental health forums. 791 forum users participated in a survey which was compared to the demographics of people accessing NHS Talking Therapies. 20 forum users took part in interviews, which were thematically analysed for reasons for use. Finally, the top keywords in the forum posts (a corpus of 28 million words) were calculated using the log likelihood test of statistical significance. One keyword was identified consistently across the forums, so its collocation profile was analysed.

Surveys showed the forums were predominantly used by white, female, and younger people, though there was greater ethnic diversity and a higher proportion of non-binary people compared to NHS Talking Therapies. Thematic analysis of interviews indicated that people used forums because they are easily accessible, making it possible to overcome barriers to in-person support, such as the need to speak. Participants sought emotional support, advice, and connection with others experiencing similar challenges. The linguistic analysis showed “scared” was a common keyword across the forums, with common collocations being *I’m scared because* and *scared of*. Reviewing the posts showed users tended to share fears over mental health symptoms and identity.

Online forums serve as important alternative and complementary sources of mental health support, particularly for people who face discrimination or logistical barriers in accessing traditional services. These forums provide accessible, anonymous spaces for peer support, helping users to share fears and connect with others in similar situations.

## Introduction

Currently, most people in the UK experience barriers to accessing mental health services due to long waiting lists with the National Health Service (NHS), as well as stricter eligibility criteria for adult services (1). NHS Talking Therapies is a state-funded scheme which provides free therapy for people in the UK. Individuals can self-refer without needing a diagnosis or to see their general practitioner (GP). It was launched in 2008 (2), and sixteen years on, the service faces immense pressure. In Southport (a UK region with the longest waiting times) people waited an average of 229 days from referral to first treatment with Talking Therapies in 2021/2022 (3). Of the 1.81 million Talking Therapies referrals received across England in 2021/22, only 1.24 million entered treatment while 688,000 completed treatment (3). The high drop-out rate in mental health services is often because of dissatisfaction with long wait times, with people who tried to use the service reporting feeling “abandoned” and “hopeless” (4). In the current climate, where demand for mental health support far outpaces the NHS’s capacity (5), online forums could provide a more accessible support mechanism.

Increasing numbers of people are accessing online forums relating to mental health (6, 7). For example, the subreddit r/mentalhealth grew from 20,000 members in 2016 (8, 9) to 530,000 in mid-2025. Alongside this, health services have a growing interest in internet-based services, with online interventions being incorporated into policy (10). The UK National Health Service (NHS) website signposts adults with mental health problems to try online forums (11), while multiple NHS Talking Therapies services sub-contracted the online forum Togetherall (previously “Big White Wall”) (12–14).

Previous research suggests that people from stigmatized groups, who experience barriers to accessing in-person services, may use forums as an alternative source of support. For example, many women experiencing perinatal mental illness do not access health services (15) because they fear judgement and the involvement of social services (16). Online forums can offer an anonymous space for peer support, which may in turn encourage people to access services, such as going to their GP (17). Similarly, people living in rural areas often experience limited access to mental health services, fear of stigmatization, and social isolation (18), which online forums can address (19).

Previous research also suggests that young people are likely to seek help online (20). Young people are particularly affected by the inaccessibility of mental health services because they are likely to have higher mental health needs (21), and less likely to be able to afford private services (22). Furthermore, as “digital natives” (23), using online search engines to find information on health issues is often their first choice (20). Social media sites like TikTok, Instagram and YouTube are increasingly used to find health-related information (24). Meanwhile, online forums remain popular, despite being one of the oldest tools for sharing information online. A systematic review (25) of 21 studies of young people’s use of online forums found that forums can provide a pragmatic resource for informational and emotional support. Other research also suggests that adolescents form the main userbase of forums to discuss eating disorders, self-harm, and suicide (26–28), which have been associated with both harmful and beneficial impacts (29).

Overall, previous research has suggested a variety of people are using different forums for different reasons. However, previous work has tended to focus on one population, in one specific forum, and collected one type of data (30). To understand who uses forums and why, it is necessary to study a range of forums, and triangulate findings across multiple types of data, each of which has methodological benefits and drawbacks. Surveys of forum users offer the opportunity for large scale and low-cost data collection but cannot offer the depth of understanding from more resource intensive interviews with users. Both surveys and interviews offer a reflexive perspective from forum users, which may be influenced by demand characteristics, memory, or social desirability bias. However, both methods benefit from being able to include people who use forums but never post, who are estimated to make up approximately 90% of forum populations (31). In contrast, corpus linguistic analysis, which has increased in use in forum research over recent years (30) can offer direct insights into what people are posting in online forums, but cannot tell us much about the people who never post. Additionally, post analysis raises ethical issues about the use of online data (32) such as whether forum posts can be considered public or private, and whether users need to consent to research (33).

Therefore, we approached the questions of “who uses online forums for mental health support, and why?”, using a novel approach of triangulating findings from multiple methods used across several online mental health forums. This study is part of the Improving Peer Online Forums (iPOF) project, which is a large interdisciplinary NIHR-funded realist-informed evaluation (34). Following a realist framework, the iPOF project aims to understand the underlying mechanisms that shape forum use: how forums work for different people across different contexts. A detailed protocol for the whole project is published (34). This study focused on the question “Who uses online forums for mental health support, and why?”, which was operationalized as the following research questions:

Who uses online forums for mental health support?

- What are the demographics, (age, gender, ethnicity) of people who use different mental health forums?
- How do the demographic and clinical characteristics of those using forums differ from the demographics of people accessing IAPT services?

Why do people use online forums for mental health support?

- What reasons do people give for forum use in a survey?
- How do people explain their reasons for using forums when interviewed?
- What are the distinctive words and phrases people use in their forum posts?

## Methods

This study used a mixed-methods approach including descriptive analysis and comparison tests with survey data, thematic analysis of interview data, and corpus linguistic analysis of forum post data. The findings from each approach are considered together in the discussion and not related to a wider realist framework.

### Ethics

Ethical approval for the iPOF project (including this study) was granted by Solihull Research Ethics Committee (IRAS 314029).

The collection of forum post data needs to be carefully considered due to ethical issues over anonymity, consent, the right for participants to withdraw, and the reasonable expectation of privacy for forum users (32). Considering these issues, we worked with forum hosts, users and moderators to develop an ethical framework for this study. To protect our participants and forum partners’ anonymity, all participants have been anonymized, and the forums have been pseudonymized using bird names. Additionally, all examples quoted from the forums are paraphrased so that they cannot be linked directly back to the forum or user.

The full ethics framework can be read online here: https://www.lancaster.ac.uk/ipof/ethics-framework/.

### Data Sources - Forums

Forum owners were approached and included in iPOF if they were UK-based (though see Discussion on the geographical diversity of the forum participants) and were focused on mental health. A multiple case series design was used to recruit seven forums, purposively sampled for diversity across hosts, target populations, design (including level and nature of moderation, and whether they require registered login), and size. Seven forums were represented across two methods of research (surveys and interviews), while the linguistic analysis of the posts was based on three forums (see details below). A brief description of each forum and the type of data analyzed from each is summarized in table 1. A full description of each forum involved in the study can be read here: www.lancaster.ac.uk/ipof/case-summaries.

**Table 1:** Brief description of each forum and the data collected.

| Table 1: Brief description of each forum and the data collected. |  |  |  |  |  |  |
| --- | --- | --- | --- | --- | --- | --- |
| Forum | Description of forum | Survey N | Interview N | Language data N |  |  |
|  |  |  |  | Words | Posts | Users |
| Sparrow | Private forum with subforums to discuss different mental health issues. | 107 | 3 | n/a | n/a | n/a |
| Robin | Private forum for people with bipolar disorder and carers. | 12 | 2 | n/a | n/a | n/a |
| Magpie 1,2,3 4, | Public forum with subforums ran by charities for different health issues. Four subforums were included in the study: Magpie.1 is about mental health issues generally; Magpie.2 provides resources and a place for discussion for people with eating disorders; Magpie.3 is for people with post-natal illness, and Magpie.4 is for people with postpartum psychosis. | 110 | 4 | 9,801,121 | 99,520 | 7,517 |
| Dunnock | Private forum for young people to discuss mental health. | 287 | 5 | 13,425,212 | 213,125 | 42,648 |
| Jay | Private forum for adults to discuss mental health. | 13 | 2 | n/a | n/a | n/a |
| Chaffinch | Public forum for young people to discuss mental health. | 49 | 1 | n/a | n/a | n/a |
| Starling | Public forum to discuss mental health generally. | 213 | 3 | 4,820,253 | 46,868 | 6,258 |
| Total |  | 791 | 20 | 28,048,586 | 359,513 | 56,423 |

### Survey

#### Recruitment

The online survey was hosted on the Research Electronic Data Capture system (REDCap, (35)). Participants were recruited from seven online mental health forums, using advertisements posted to the forum and, when possible, through direct emailing to registered forum users. Participants were included if they were a) over 16 years old, b) a UK resident, and c) had visited the forum at least once before, which were all measured through self-report. After passing the inclusion criteria, participants were emailed a unique link to complete the survey. The survey included questions on the participants’ engagement with and views of the forum, their mental health, their engagement with other health services, and their demographic information. Participants received a £10 digital voucher upon completion. The final dataset consisted of 791 survey participants, from seven forums.

#### Materials

Data from specific questions of the survey were used. Demographics (age, gender, ethnicity) were measured using categorical response options. The reasons for joining the forum were recorded with multiple response options, as well as the opportunity to write an answer. An example is displayed in Textbox 1, while all survey questions used in this study can be found in Supporting Information 1.

##### Textbox 1. Example survey question on the reasons for forum use

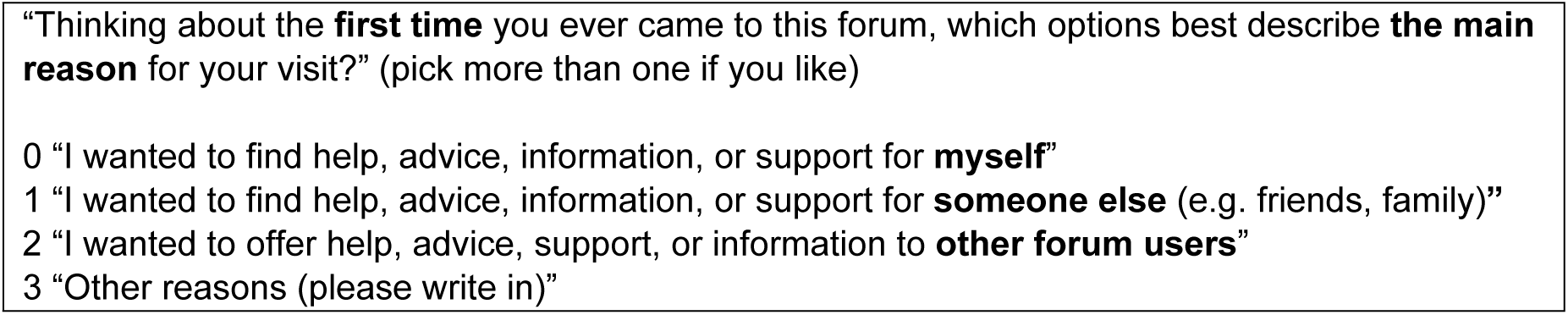

### Comparison Dataset: Talking Therapies

Demographic data from the 1,215,329 people accessing NHS Talking Therapies between 2022-23 was used to compare with the survey’s demographics. This data was sourced from the publicly available NHS Talking Therapies Annual Report 2022-23 (36).

### Statistical analysis

Data management and analysis were conducted using Microsoft Excel and SPSS.

The frequencies were reported for the age, gender, and ethnicity of participants from each of the seven forums. The survey results were recoded to ensure comparability with Talking Therapies (with details in Supporting Information 1). To compare the demographics of the forum users with the demographics of people accessing Talking Therapies, chi-squared comparison tests were conducted. The whole dataset of 791 participants from across the seven forums was considered as one “forum-user” group.

### Interviews

#### Recruitment

A convenience sample of twenty forum users were recruited through forum advertisements and through expressions of interest on the survey. Interviews could take up to an hour, and participants received a £30 digital voucher as a thank-you for their time.

#### Materials

Interviews were conducted remotely, using Microsoft Teams for video calls or phone calls depending on the participant’s preference. Interviews were recorded using an encrypted voice recorder and the recording was deleted after transcription. Transcription was conducted by an independent transcriber.

Semi-structured realist-informed interviews were conducted by PM and ZG to openly explore participants’ experiences of using forums, before progressing to more specific testing of theories on how online forums work (37). The full topic guide is available in Supporting Information 2. The analysis for this study focused on participants’ responses to the first section of open-ended questions, including “what motivated you to start using it?”.

A reflexive thematic analysis, following the guidance of Braun and Clarke (2006; 2019) was conducted, exploring why participants used the forums. The analysis process began with data familiarization; ZG read all twenty transcripts. Interviews were coded inductively by ZG for reasons for forum use, using NVivo (38). Preliminary coding and themes were discussed by ZG, FL and PM. After these discussions, ZG refined the coding and thematic structure, and later iterations were discussed in a meeting with ZG, FL, PM, ES, and PR.

### Linguistic Analysis of the Forum Posts

#### Data Collection

The language data was collected from three forums, Starling, Dunnock, and Magpie. Magpie included four subforums, which were kept separate for this analysis.

The three forums were selected to represent different user demographics and different hosts, which ensured a diverse range of contexts were included, while also keeping the full dataset a manageable size by not including every forum.

A nuanced data collection approach was used for the different forums. For Starling, a public forum, we worked with the hosts and moderators and gave users the opportunity to opt-out of the research before scraping the forums. For Dunnock and Magpie, the forum hosts shared the post data from consenting users with us.

#### Analysis

We used a combination of both quantitative and qualitative methods to identify linguistic patterns that are relevant to our research questions. As our research questions were broad, we first adopted a well-established corpus linguistic technique known as ‘keyness’ analysis. This involves comparing the relative frequencies of words in a ‘target’ corpus with the relative frequencies of words in a ‘reference’ corpus. The output is a list of words, known as ‘keywords’ that are statistically significantly more frequent in the target corpus than in the reference corpus. In our analysis, each of the six forums mentioned above was treated as a target corpus: Starling, Dunnock and the four Magpie forums. A corpus sampled from the written subcomponent of the 1994 version of the British National Corpus was used as a reference corpus (39). We used the log likelihood measure of statistical significance (40) to calculate and rank the keywords from each of the forums. We compared the top ten keywords in each of the target corpora (six keyword lists altogether) and their log likelihood values. In the comparison, we aimed to identify keywords that occurred consistently across the top ten keywords in all the keyword lists. The comparison showed there is only one such keyword (*scared*) and this was further analyzed through its collocation profile. In corpus linguistics, collocates are words that co-occur in close vicinity next to each other in a statistically significant way and they reveal important information on the word’s linguistic usage (41). The collocation analysis was performed in AntConc software (42). To calculate the collocations, we used likelihood statistics and a span of five words to the left and right of the keyword. Our dataset is, in terms of its size, heavily skewed towards posts retrieved from Dunnock – 48% of the total word count, while posts from Starling take up only 17% of the overall wordcount. For the collocation analysis, we have therefore added an additional condition for the collocate to occur in all three forums to be considered for detailed analysis.

## Results

### Survey

As shown in Table 3, across all the forums, most survey participants were white and female, and the most common age bracket was 16-24.

**Table 3:** Demographics of the forum survey.

| Table 3: Demographics of the forum survey |  |  |
| --- | --- | --- |
| Demographics |  |  |
| Age | N | % |
| 16-24 | 285 | 36 |
| 25-34 | 196 | 24.8 |
| 35-44 | 133 | 16.8 |
| 45-54 | 68 | 8.6 |
| 55-64 | 65 | 8.2 |
| 65+ | 41 | 5.1 |
| Prefer not to say | 3 |  |
| Gender | N |  |
| Female | 474 | 59.9 |
| Male | 253 | 31.9 |
| Non-binary | 46 | 5.3 |
| Prefer not to say | 13 | 1.6 |
| Prefer to self-describe | 5 | 0.6 |
| Ethnicity | N |  |
| White | 643 | 81.3 |
| Black | 37 | 4.7 |
| Asian | 35 | 4.4 |
| Mixed | 55 | 6.9 |
| Prefer not to say | 13 | 1.6 |
| Prefer to self-describe | 8 | 1.01 |

### Comparison with Talking Therapies

Table 4 shows the comparison between the survey group and the Talking Therapies group. Significant differences were found across each of the sociodemographic variables, although the effect sizes were weak (43). The comparison suggests a greater proportion of young people aged 16 – 25 accessing online forums, compared to people accessing Talking Therapies. There was a smaller proportion of females accessing online forums compared with Talking Therapies, despite females being the dominant gender across both datasets. There was a larger proportion of the white and mixed ethnic groups accessing online forums compared with Talking Therapies, although the white group was the majority across both datasets.

**Table 4:** Chi-squared comparisons between forum and Talking Therapies samples.

|  |  |  |  |  |  |
| --- | --- | --- | --- | --- | --- |
|  | iPOF Forums<br>n=791 |  | Talking Therapies<br>n=1,215,329 |  | Chi-Squared Test |
|  | % | 95% CI | % | 95% CI |  |
| **Age** | | | | | $\chi^2(3)=4703.21$,<br>$p<.001$,<br>Cramer's<br>$V=0.062$ |
| Under 25 | 36.0 | (32.7 – 39.4) | 21.9 | (21.8 – 22.0) |  |
| 25 – 64 | 58.4 | (55.0 – 61.8) | 71.3 | (71.3 – 71.4) |  |
| 65 and over | 5.2 | (3.6 – 6.7) | 6.8 | (6.7 – 6.8) |  |
| Not stated | 0.4 | (0.0 – 0.8) | 0.0 | 0 |  |
| **Gender** | | | | | $\chi^2(3)=856.00$,<br>$p<.001$,<br>Cramer's<br>$V=0.027$ |
| Male | 32.0 | (28.7 – 35.2) | 31.8 | (31.7 – 31.8) |  |
| Female | 59.9 | (56.5 – 63.3) | 66.5 | (66.4 – 66.6) |  |
| Indeterminate | 6.5 | (4.7 – 8.2) | 0.3 | (0.3 – 0.4) |  |
| Not stated | 1.6 | (0.7 – 2.5) | 1.4 | (1.3 – 1.4) |  |
| **Ethnicity** | | | | | $\chi^2(5)=84.19$,<br>$p<.001$,<br>Cramer's<br>$V=0.008$ |
| Asian | 4.4 | (3.0 – 5.9) | 6.8 | (6.8 – 6.9) |  |
| Black | 4.7 | (3.2 – 6.2) | 4.1 | (4.1 – 4.1) |  |
| Mixed | 7.0 | (5.1 – 8.7) | 3.2 | (3.2 – 3.3) |  |
| White | 81.3 | (78.6 – 84.0) | 76.2 | (76.2 – 76.3) |  |
| Other Ethnic Group | 1.0 | (0.3 – 1.7) | 2.3 | (2.3 – 2.4) |  |
| Not stated | 1.6 | (0.8 – 2.5) | 7.3 | (7.2 – 7.3) |  |

### Breakdown across the forums

As shown in figures 1, 2 and 3, there was some variation across the different forums in the participant demographics. Dunnock and Chaffinch (two forums aimed at young people) had the highest proportions of young participants, with 72% (241) of their participants in the 16-24 bracket. In contrast, 89% (98) of Magpie (a public platform with charity-run sub-forums) participants were aged 45 and over. Sparrow (an NHS based forum) had a slight majority (28%, n=30) of participants in the 35-44 bracket, although there was representation of participants across all age brackets. Participants from 6 out of 7 forums were majority female. In Starling (a public forum to discuss mental health generally), the majority was male. Chaffinch showed the highest proportion of non-binary participants (22%, 11), which was the second most frequently reported category after female. Participants were mostly white across all forums.

**Fig. 1:**
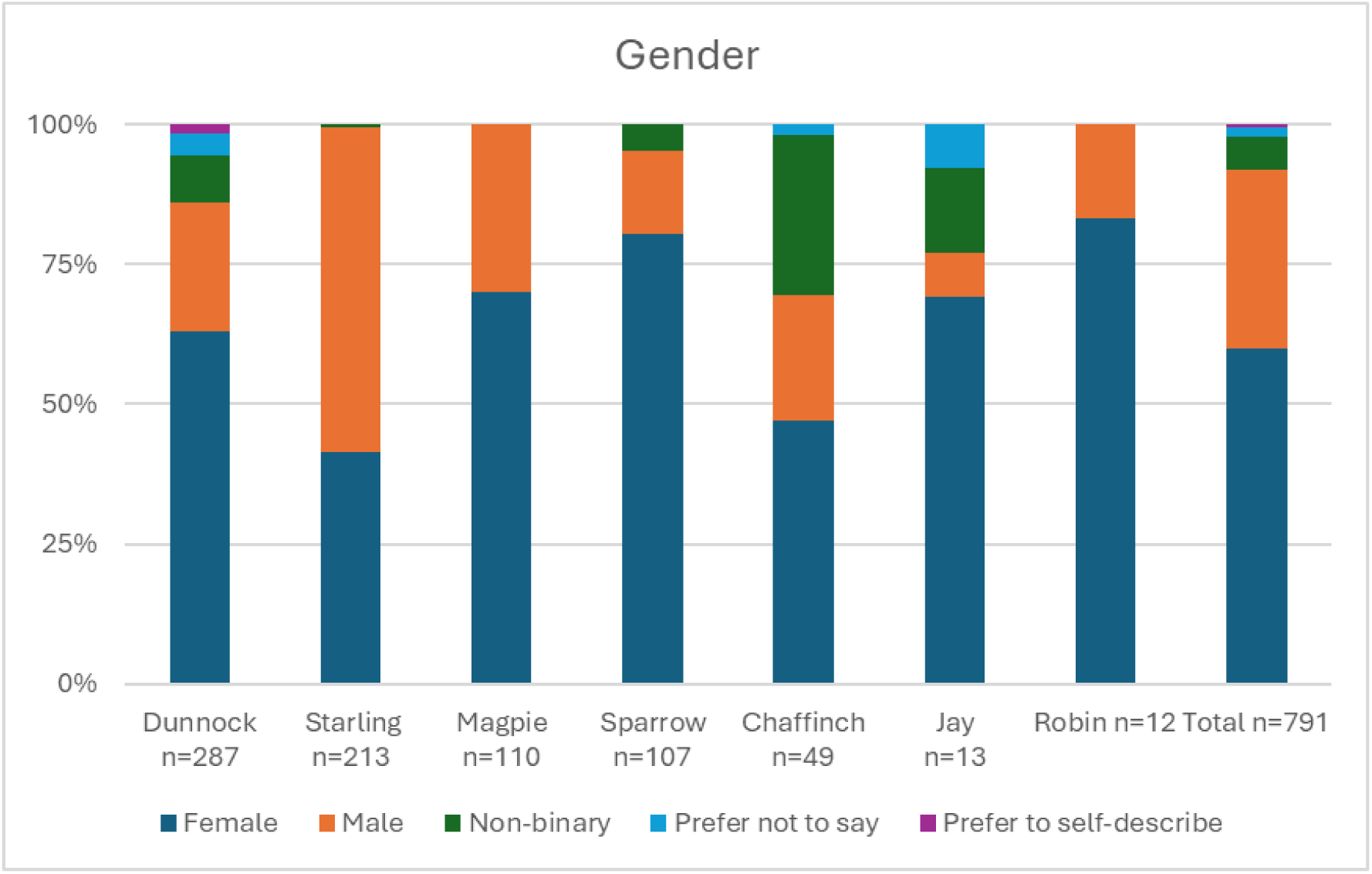
Breakdown of gender reported on survey

**Fig. 2:**
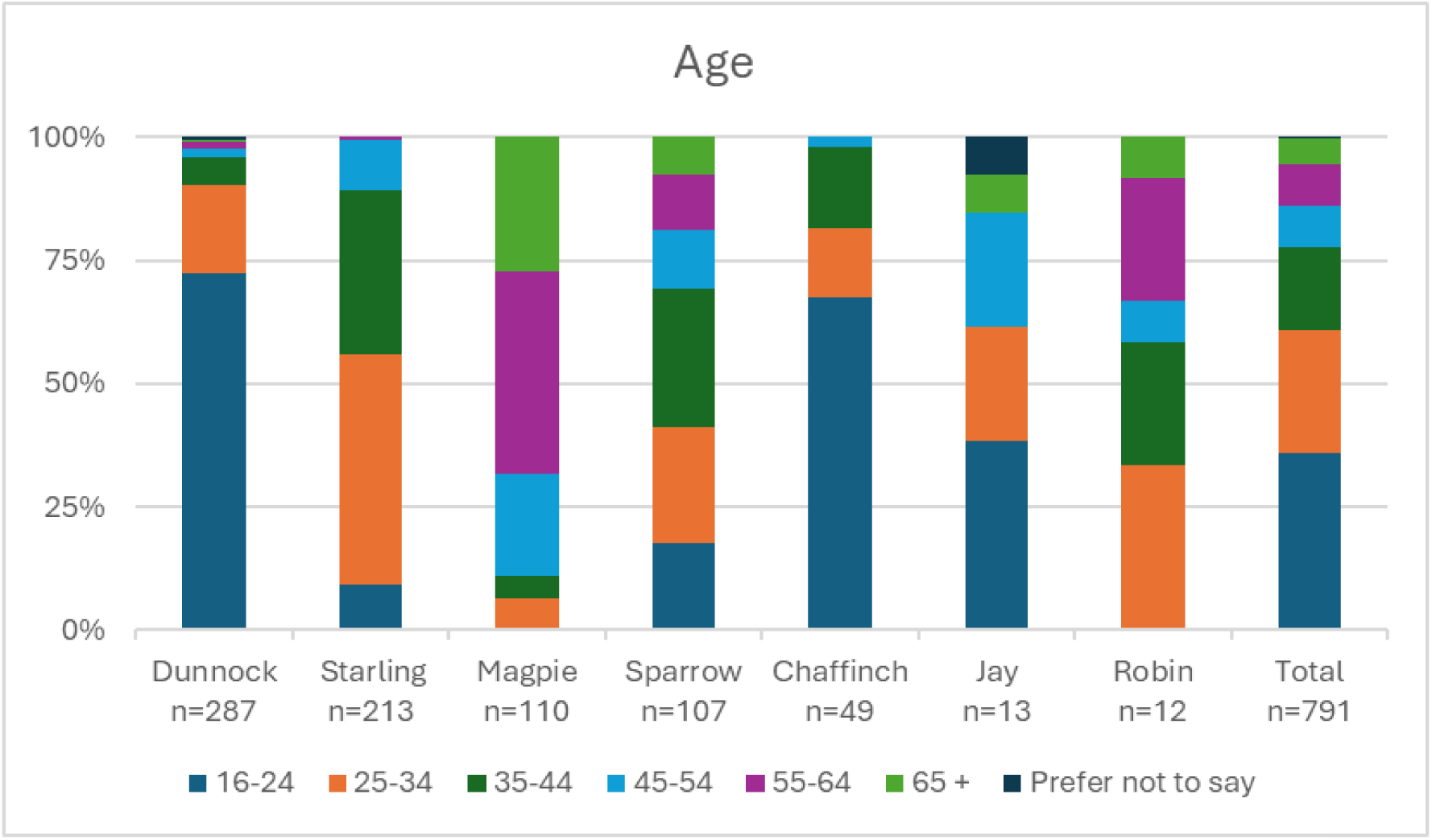
Breakdown of age reported on survey

**Fig. 3:**
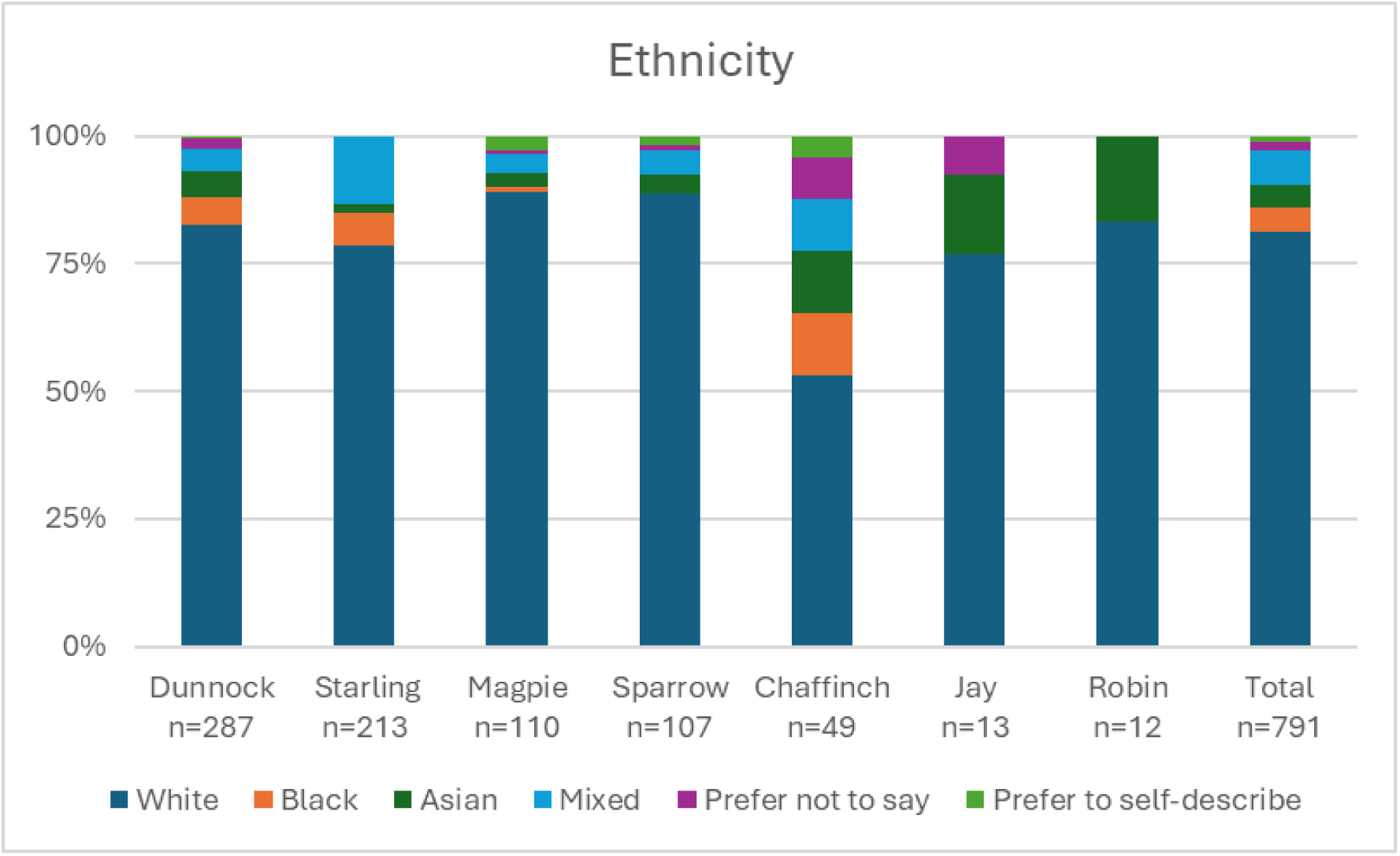
Breakdown of ethnicity reported on survey.

### Reasons for Use

Table 5 shows the proportion of reasons that respondents gave for visiting the forum.

**Table 5:**

| **Table 5: Reason for visiting forum** | **N (%)** |
| --- | --- |
| I wanted to find help, advice, information, or support for **myself** | 544 (69%) |
| I wanted to find help, advice, information, or support for **someone else** (e.g. friends, family) | 237 (30%) |
| I wanted to offer help, advice, support, or information to **other forum users** | 229 (29%) |
| Other reasons | 20 (3%) |

Twenty participants mentioned other reasons, including wanting “to vent to others”, wanting “to see if other people had been through similar things so I didn’t feel alone”, as well as because the forum was recommended to them by others.

### Thematic Analysis of Interviews

The sample of 20 participants were mainly White/White British, female and in the 16-25 age bracket. Full demographic data is displayed in Table 6.

**Table 6:**
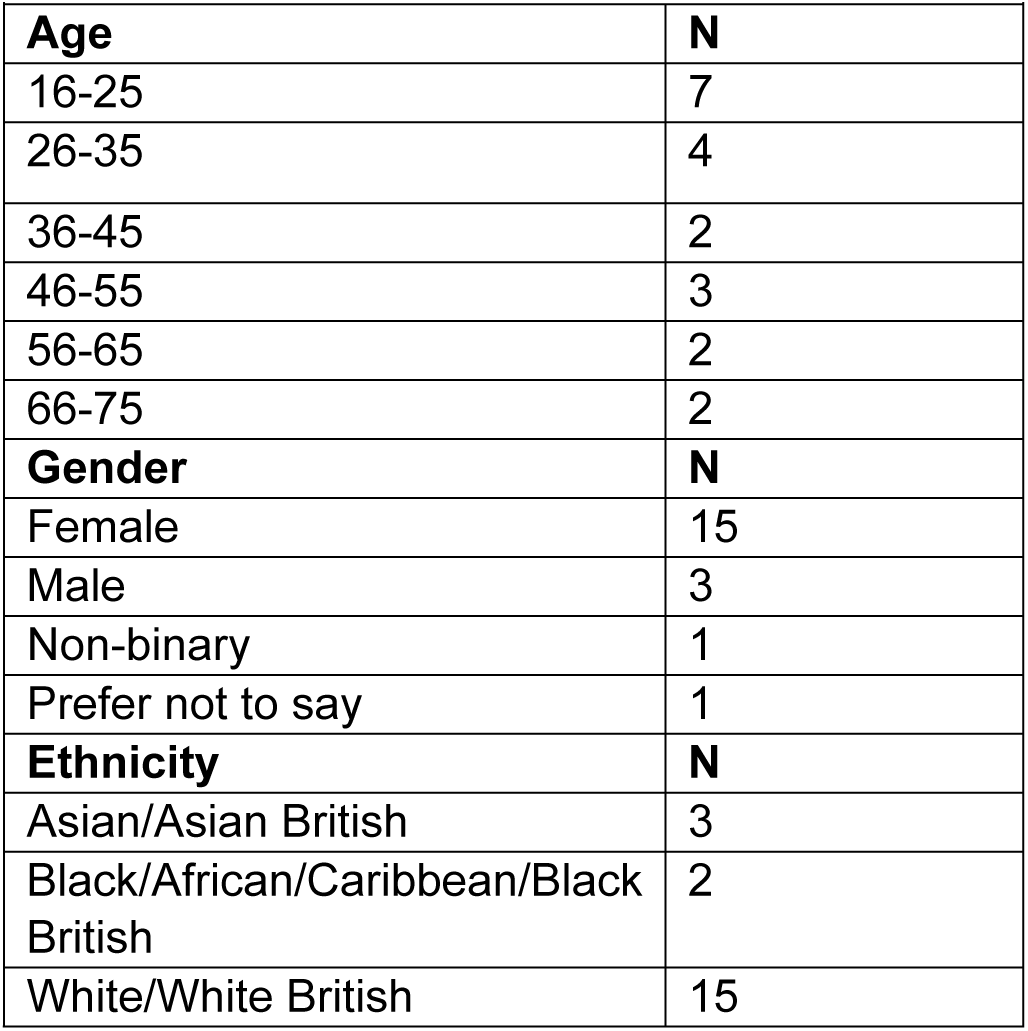
Demographics of interview participants.

| **Table 6: Demographics of interview participants** |  |
| --- | --- |
| **Age** | **N** |
| 16-25 | 7 |
| 26-35 | 4 |
| 36-45 | 2 |
| 46-55 | 3 |
| 56-65 | 2 |
| 66-75 | 2 |
| **Gender** | **N** |
| Female | 15 |
| Male | 3 |
| Non-binary | 1 |
| Prefer not to say | 1 |
| **Ethnicity** | **N** |
| Asian/Asian British | 3 |
| Black/African/Caribbean/Black British | 2 |
| White/White British | 15 |

Thematic analysis of the interviews suggested some key reasons for using forums, including forums’ availability and asynchronous written nature. These may be especially important for young people who have experienced the lockdown-induced shift to online communication.

Some participants used forums to complement their in-person social and clinical support, whereas others reported joining forums as an alternative because they could not access NHS services or because they could not find offline peer support. Some participants had stopped using forums because they could not find the help, advice, or information they sought. However, others stayed and kept using the forum because of the opportunity to help others. Illustrative quotes are included in each theme, with more quotes available in Supporting Information 3.

#### Anytime, Anywhere: The Value of Accessibility

People often first find forums after searching the internet for specific experiences, opinions, or advice. The public nature of forums such as Starling or Chaffinch means that discussion threads can appear in Google search results. Easily accessible public forums can provide information, advice, or a sense of shared experience, for people even if they do not register or create any posts. Similarly, on closed forums, such as Dunnock and Robin, members can use the search feature to look for previous posts on specific experiences, which was often preferred over creating an original post.

> “*if I’m having issues then I’m not sure but I might want to actually see if there’s any posts on it beforehand – I prefer not to actually have to ask or create my posts. I prefer to do that and yeah so try and look through if there’s any posts similar to my issue so yeah I just kind of look through those*”. Starling user.

Unlike other modes of online peer support, such as Zoom support groups or one-to-one webchats where conversations are transient, the conversations posted to forums are often permanently and publicly available for others. Therefore, supportive posts sharing experiences or advice between users on the forum go beyond one-to-one peer support, because the content is and will always be accessible for others searching for it, helping an unknown number of future viewers.

Online forums are easily accessible on a day-to-day basis. For most forums, account creation takes minutes, requiring a valid email and the creation of an anonymous username. Being able to access the forum on a mobile phone which can store log-in details was important for users to easily read posts or connect with others at any time of the day, anywhere.

> “*You can make an account very quickly and you can just join in just like that*”. Chaffinch user.

> “*If it wasn’t on my phone I don’t think I’d use it. Literally I’m permanently logged in on the memory – so I just type [Sparrow] and it comes up with the username and log in … I don’t think I would specifically open my laptop to go on it*”. Sparrow user.

Forums can therefore offer interpersonal connections, mental health advice and support at any time, in between more structured support, such as therapy appointments or scheduled peer support groups.

> “*it was advised – or it was offered to me by a cognitive behaviour therapist counsellor as an extra resource in the week when I was speaking to her*”. Sparrow user.

The 24/7 availability of forums may be especially important for people with specific mental health issues. For example, Robin had a “can’t sleep” thread, allowing users to support each other when awake at night. There are other online options available 24/7 for mental health support, such as helplines or one-to-one webchat, but these can have waiting times and are often targeted to people experiencing acute distress, such as feeling suicidal.

> “*Sometimes you feel like it’s dark and everybody is asleep and you’re just alone suffering by yourself but being able to access the forum and this particular thread it makes you feel a bit nicer*”. Robin user.

> “*You can call Samaritans at three in the morning but you might not want to. We do often have users and I’ll be on late at night if I’m not sleeping so you can kind of get 24 hour support… You don’t have to leave the house to access the forum. You don’t have to talk to anyone*”. Starling user.

However, the ability to access forums at any time can be detrimental to the user, which may also become a reason why people leave. For example, while users struggling to sleep may feel supported by others, screen time at night could ultimately disrupt sleeping patterns further. Similarly, unrestricted access to a forum about anxiety could contribute to a person’s rumination and ultimately worsen their anxiety. These concerns may be especially pertinent for forums which are more like social media, such as Starling and Magpie, which increase user engagement though storing log-in details, sending post notifications via email or app, and creating an endless algorithmically curated feed of posts for users to scroll through. These strategies push users to engage passively with the forum. Posts on mental health forums inherently contain some discussion of distress, and pushing users to consume this content may become detrimental to their wellbeing.

> “*I feel like I reflect more on my own anxiety and I sort of increase my anxiety because of that so I think that’s probably where my problem lies with that sort of community… To be honest, I think the only one [forum] that would probably have a negative impact on my mental health is a mental health forum.*”. Magpie user

#### “Happier to Type”: In Your Own Time, Space and Words

Forums are not only used because they are easily accessible, but also because of the written nature of communication. Typing and reading, rather than speaking, creates a sense of psychological distance and safety, and can have therapeutic benefits.

> “*it’s not like meeting somebody or phoning somebody where you have to have an immediate answer*”. Sparrow user.

Firstly, reading and responding to posts is an asynchronous process; there is no pressure to respond in real-time to other users. This is important to the appeal of forums for people with communication difficulties, such as being deaf, being neurodivergent, or having severe anxiety. For these groups, spoken communication, ranging from telephone or in-person conversations with health professionals to chats in informal social settings, can be challenging because of the pressure to understand and respond quickly, or the need for an interpreter. Forums therefore can provide a way of connecting to others in one’s own time, without the chance of mishearing, and without needing to communicate through third parties such as an interpreter.

> “*it’s so much easier to be able to just read and type… it’s all just texting and reading regardless of whether you have issues with hearing, lipreading or anxiety with speaking when you’re really distressed or autism*”. Jay user.

As well as being asynchronous, the written nature of forums also creates a sense of anonymity and ease that cannot be achieved through other online services, such as helplines, talking therapies, or groups, where individuals would have to speak. People with mental health issues can struggle to verbally speak about their issues, especially if they are distressed or under pressure which can elevate heart rate, tighten the throat, and generate racing thoughts, all of which make it harder to speak.

> “*I had counselling and stuff before but I wasn’t – it sounds really weird but I wasn’t in a position where I wanted to talk. I was happier to type, so I needed somewhere I could go where I didn’t have to verbalize what was going on, I could write it down*.” Dunnock user.

> “*I wouldn’t imagine many people whether they’re autistic or not when they’re crying or shaking when they’re really stressed actually wanting to <u>talk</u> talk*” Jay user.

This may be especially important for younger people who experienced the lockdown-accelerated shift to online communication at an earlier age. Online communication is the norm for this group, meaning in-person talking counselling may feel uncomfortable.

> “*Lockdown had a huge impact especially on my generation I’d say…because we’re all so used to talking online. I think when I went back into school I found it especially stressful. Talking to people face to face again was weird… I think that deterred me from talking to people as well after lockdown the fact that I’d just grown used to it online*”. Dunnock user.

Like journalling, typing online can also encourage individuals to self-reflect, express their feelings, and track progress over time across multiple posts. If users are actively engaged with the forum and taking time to write and reflect, typing can be a cathartic process. Furthermore, the medium of forum post expands on journal entries because it allows other people to read it and is effectively posted permanently. The visibility of forum posts can serve multiple purposes: it allows others to share advice; it allows the user to feel “seen”, and it can help the user externalize their own experiences and see the content from a new perspective.

> “*yeah it does help, it just externalizes it from yourself. You’re sort of almost then looking in on it from other people’s point of view and you also get support which is helpful*” Sparrow user.

#### “I’ve got nobody else to ask”: A Last Resort

Limited access or poor quality of NHS services is another reason why people join forums, as an alternative source of advice and information. This is different to the reasons discussed previously in the “accessibility” theme, where people use forums as a complement to other services, such as by accessing the forum on their psychiatrist’s advice or between therapy sessions. In contrast, some people try forums as a complete alternative because they cannot access mental health services. This could be due to long NHS waiting lists, a lack of support after diagnosis, or inadequate treatment options.

> “*What started me using them is the complete lack of information about my mental health conditions that I received from the NHS. I was given a bipolar disorder diagnosis and not given any guidance, not even a leaflet about what it was or what you had to do about it…. so the main reason I’ve been looking into forums is because I’ve got nobody else to ask.*” Starling user.

> “*I’ve had a particularly difficult – I’m still in the throes of a particularly difficult time of my life. Quite a lot happened all in one go. I’ve really struggled to access decent support from my GP which is what I really needed at the time and I googled a way to get some help and be able to talk to someone*” Dunnock user.

While for some people, forums can provide a helpful alternative source of advice and support, for others it only exacerbates their feelings of distress, isolation, and disenchantment with the health system. This may depend on the severity of mental health issues faced by the individual. It may also be because these individuals are trying forums as a last resort and would prefer spoken communication with health professionals instead, in contrast to the kinds of people described in the previous section.

> “*I got to the point with a therapist on [Dunnock] she was doing my head in. she was absolutely useless and not helpful and fairly judgmental and I got to the point where it was actually doing me more harm than good using [Dunnock] and obviously I tried again with my GP to get some support and help and they were more than useless so I found a private trauma counsellor… that’s a bit more sustainable and sensible*”. Dunnock user.

#### Finding People Like Me: Social Connection

Forums were used to connect socially with people with similar experiences or identities, who might not be accessible in person. Even for those who have an offline friendship group, they may not feel comfortable sharing their mental health issues because they fear being misunderstood. The forum can provide a way to initiate and build friendships with people who understand the individual’s perspective. This may be especially important for people with disabilities that limit their ability to leave the house – an online forum can provide a lifeline to connect with others with similar experiences.

> “*I have a very supportive social circle but there have been moments where I have felt isolated or I have isolated myself from my social circle because I felt like they didn’t understand what I was going through… so when I see other people going through similar situations I’m motivated to talk to them*”. Chaffinch user.

> “*because I’m nonbinary I think I kind of appreciate the trans nonbinary people’s responses sometimes because if it’s a situation to do with how I experience dysphoria then I think sometimes that’s something that only other trans nonbinary people actually do experience in the same way*”. Jay user.

Some individuals, however, may still struggle to find others online with similar experiences, especially if they feel their experiences are uniquely challenging, and this can again become a reason why users become frustrated and stop using forums.

> “*I would say [I’ve tried] maybe twenty odd forums and I find that they all kind of say the same sort of thing and I think – I know sometimes it’s changed but I think my situation is a lot more chronic than a lot of people there because they – I’ve got a lot of issues going on so my problem is quite unique I think, so I can’t resonate with – I look at people that are in them because they’ve got maybe one issue to deal with whereas I’ve got maybe five or six so it’s difficult to try and get a rapport with people like that.*”. Magpie user.

#### Giving Back to the Community

As well as exploring why people join forums, it’s important to consider why people keep returning to forums too. While many users will never return to the forum at a certain point, either after finding the help they need or becoming frustrated by the lack of it, others keep revisiting the forum even when they are no longer seeking help themselves. With more time on the forum, and more time and experience living/recovering with mental health issues, the role of the user can shift from being a support-seeker to a support-giver, and even a moderator. This transformation is driven by a desire to help others in similar situations and give back to the community, either by sharing their lived experience, providing advice or comfort, or simply letting others know that they are there to listen.

> “*back when I was very unwell I used to post a lot and ask a lot of advice and share my feelings and talk to people about my feelings but now I’m feeling better and in a more stable position I tend to use the forums so I can help others and it gives me a real sense of achievement and having some use that even though it’s a stranger that I don’t know, I’ve never met and will never meet just by reaching out to them and going, ‘I’ve been in that position. This is what I did.’ I feel like I make a difference for other people and it makes me feel less useless because I feel if I can help somebody who hasn’t got help like me then there’s still a point in me being here.*”. Starling user.

### Linguistic Analysis of Forum Posts

#### Keywords

Table 7 shows the top ten keywords from each forum corpora.

**Table 7:** Top ten keywords in each forum.

| Table 7: Top ten keywords in each forum. |  |
| --- | --- |
| Forum | Keywords |
| Magpie.1 | *Hi, **scared**, nt\*, mg, depression, medication, anymore, none, hugs, mas\** |
| Magpie.2 | ***scared**, disorders, disorder, calories, hi, referral, nt, scary, ed\*, helpline* |
| Magpie.3 | *Hi, symptoms, **scared**, symptom, pregnancy, yrs, horrendous, anymore, debilitating, mums* |
| Magpie.4 | *Hi, medication, mums, pregnancy, lithium, **scared**, scary, delusions, mg, diagnosis* |
| Dunnock | *nt, **scared**, hi, anymore, ur, scary, bisexual, honestly, bullied, self* |
| Starling | *diagnosis, referral, mental, therapy, medication, therapies, suicidal, trauma, **scared**, disorders* |

\* nt = not, mas/ed = forum related abbreviations

The word *scared* is, based on the log likelihood statistics, the only top 10 keyword that is shared across the six datasets. The word *scared* itself occurs in the full dataset over 10,300 times, being relatively (in relation to the dataset size) the most frequent in Dunnock. As Dunnock is, at the same time, also the biggest dataset, the following collocation analysis needs to be interpreted as primarily representing the contributions made by Dunnock users.

| Forum | Total word count | **scared** occurrences |
| --- | --- | --- |
| Dunnock | 13,425,212 (48%) | 7,368 |
| Magpie | 9,801,121(35%) | 2,332 |
| Starling | 4,820,253 (17%) | 679 |
| TOTAL | 28,048,586 | 10,379 |

### Collocates of “scared”

We calculated the collocates of the word *scared* to establish the major linguistic patterns of its usage across all the dataset combined. The top 60 collocates, their frequencies and log likelihood statistics values are included in Supporting Information 4; this also includes the collocates calculated for each forum separately to show the ranking and values of the collocates we analyze below.

The top collocates include many function (grammatical) words, including various alternative spellings: collocates *’m, I, im, am* all mainly occur within the phrase *I’m scared,* while prepositions and conjunctions, such as *because, of, to* indicate they may be part of phrases revealing reasons for being *scared.* We decided to look in detail at two of the top collocates: *because* and *tell. Because* (rank 4) is the first collocate that follows collocates that make up the phrase *I’m scared* (ranks 1 to 3) and *tell* is the first lexical collocate, i.e. a word with a meaning, not a grammatical word.

### Collocate “because”: I’m scared because / because I’m scared

There is a fairly even distribution between occurrences of *because* within five words on the left and right of *scared*, which suggests that contributors both give reasons why they (or someone else) is scared and present being scared as a reason for something else.

> *because I am scared [X] will judge me* [Dunnock]

> *I’m scared to tell [X] because I don’t want …* [Dunnock]

This collocational pair occurs in an overwhelming majority in Dunnock posts (only 6% of occurrences are found in Starling and 17% in Magpie). We further narrowed down the analysis to the phrase *scared because* (208 occurrences) as we hypothesized this may reveal reasons not only to why the participants are scared but also be relevant to wider reasons for them being on the forum. Here, the majority occurred in the first-person pronoun (87% of occurrences), with the majority occurring again in Dunnock (78%).

The qualitative analysis shows that participants share both their fears of their past experiences, fears of past experiences that are still present in their lives, and fears of current situations they find themselves in. The reasons for being scared seem to be similar across the forums, ranging from being scared of concrete things (e.g. spiders), to being worried about relationships, and very frequently of symptoms, the unknown and associated feelings:

> *Also I’m scared because they might give me medication.* [Dunnock]

> *I am scared because at night I start panicking and no longer feel in control …* [Magpie]

> *I’m quite scared because I think you’re not allowed to stop taking the meds, but at the same time, if I don’t have a follow up appointment before they run out, what am I supposed to?* [Starling]

In addition to these general themes, in Dunnock, we could identify several specific themes, which may arguably be linked to the forum’s participants’ younger age. These included themes of fear associated with school environment, both socially and in relation to academic pressure, themes of self-harm, themes of sexuality and “coming out” and relationships, including romantic relationships, more generally.

> *I want to do really well in all subjects and I’m especially scared because you have to do science GCSES and I’ve only got a grade 5* [Dunnock]

> *I’m scared because school makes me really anxious and this is the root to so many of my issues* [Dunnock]

> *I’ve been self-harming and I’m sort of scared because the thoughts are getting much worse, and I really don’t know what I should do* [Dunnock]

> *It will be long before we can marry but will she still want to get married? I’m just scared because I don’t want to lose her.* [Dunnock]

> *so I am gay and I want to come out to friends but im really scared because I am not sure how they will take it. does anyone have any advice…* [Dunnock]

> *So I am thinking that I might be trans and I’m scared because I have no one to talk to* [Dunnock]

### Collocate “tell”: scared to tell

The collocate *tell* (495 occurrences with *scared*) is, similarly as *because,* in an overwhelming majority of occurrences (89%) linked to the Dunnock forum; this is also because it is statistically more significant among the collocates in Dunnock. More than half of these occurrences (54%) occur in a pattern where the subject is the first-person pronoun (*I*) followed by *be* verb in various forms (most frequently *am* and *was*), with several other stylistic variations with similar meaning, such as *I felt scared*.

This pattern frequently includes references to people they are *scared to tell to.* These may be general actors, such as *anyone, someone, people* (30% of occurrences) and people who are close, such as *family*, *parents,* mother and father (39%), and/or people in a position of trust, such as *friend(s)* and *teacher(s),* but, interestingly, also *doctor, nurse, therapist, councilor, CAMHS worker.* This is similar across all the forums.

While the context of occurrences is not always sufficient to determine the causes of fear, the most frequently mentioned recurring themes are broadly similar to the ones seen in the analysis of the collocate *because,* linking to sexuality (e.g. “coming out”) (21% of instances) and mental health problems and associated experiences. The description of the causes of fear is occasionally quite complex and includes cases where participants state that they suspect they may have a diagnosis which has not been confirmed yet or are scared to get a diagnosis and seek help and advice, as shown in the examples below. The data additionally contain several examples of direct requests for advice, most frequently on “how to tell” someone of their fears or offering encouragement:

> *I need an advice! Can anyone please help because I’m scared to tell my camhs worker so that they don’t think I’m self diagnosing.* [Dunnock]

> *My parents don’t know I am gay and I am scared to tell them what do I do? Any advice anyone?* [Dunnock]

> *Don’t be scared, tell the crisis support staff how you are feeling. They are there to help you. You’ll be ok.* [Magpie]

> *But, you know, I’m really scared. Can anyone tell me what their experience was like?*[Starling]

Some of the examples explicitly mention forum participants appreciating that they can meet people they perceive as similar in the forum and that they appreciate the anonymity of the forum to freely share and find more information.

> *I have depression and [X] but I haven’t received an official diagnosis because I’m too scared to tell anyone. It’s kind of nice to see other people so similar to me.* [Dunnock]

> *I think I have [X] but I’m not quite sure. I haven’t been diagnosed because I’m too scared to tell anyone. I was hoping with the anonymity here to get some answers.* [Dunnock]

The majority of the posts that include this collocational pair (80%) employ “being scared to tell” in the present tense (*I am scared*), suggesting that forum participants are sharing experiences they are going through at the moment. However, there are posts where this collocational pair is part of the past tense structure, in several instances aiming to convey a positive message of “having told” and consequently feeling better, or even being articulated in the form of encouragement, see the examples.

> *I was terrified and scared to tell people. But now after having been open about it, I feel like I can finally breathe again!* [Dunnock]

> *Hey I have been diagnosed with [X] and was really scared to tell anyone similarly like you. I told my parents two weeks ago and they were really supportive.* [Dunnock]

> *I had a friend like this and I was scared of him but I trust in you, you should stand up to him and tell him how he makes you feel* [Dunnock]

## Discussion

This study has approached the question of who uses forums for mental health support and why from three methodological angles, across seven different forums. Firstly, the survey indicated that most participants were white, female, and in the 16-24 age bracket, although the breakdown of results showed variety across the forums. Compared to Talking Therapies, there was a smaller proportion of females in the forum dataset, and a greater proportion of people aged 16-25 and a greater proportion of people identifying as white or mixed ethnicity, although these differences were mostly small. Most survey participants (69%) reported joining the forum to find help for themselves. Secondly, the thematic analysis of interviews provided deeper insights into the key reasons why people first joined forums, as well as why they kept using them or left. These were the initial availability of the forum, the asynchronous and written nature of forums, a lack of support from the NHS, a desire to form social connections, and staying on the forum to help others. Finally, the linguistic analysis explored the keywords across three forums and then in more detail explored the one word which was in the top keywords across all the examined forums: *scared*. The linguistic analysis suggested that people were using forums to share their fears, which often were around symptoms or fears relating to mental health treatment itself, and fears around questioning gender and sexual identity. These fears were shared more easily in the online forum than in offline relationships including with friends, family and therapists.

Data from all three methodologies suggests forums may be appealing especially to young people who are transgender or questioning their gender identity, which aligns with previous research suggesting that forums are useful for people experiencing stigma and barriers to healthcare. 5.3% of the survey sample identified as non-binary, while only 0.3% of the population accessing NHS Talking Therapies were recorded as “indeterminate”. Notably, the non-binary proportion of the survey sample was mainly from the forums with younger demographics: Dunnock and Chaffinch. Our thematic analysis showed some people use forums as a last resort when they cannot access adequate support from mental health services. Previous research has suggested that transphobia is a key barrier for transgender people accessing health services (44), which has been attributed to a lack of therapists’ professional knowledge, as well as a lack of therapists with lived experience of gender dysphoria (45). The thematic analysis also highlighted the importance of forums as mediums to connect with people with shared lived experiences, which includes shared understandings of mental health experiences, and of gender dysphoria. Similarly, the linguistic analysis found that many forum users shared that they were scared of “coming out” to friends and family and were seeking advice from others on the forum. This supports previous research which has shown that transgender youth value online forums because of the support networks they provide as well as practical information (46).

Another finding that emerged across the methods was that a subgroup of people use forums to help other people who were going through the same experiences that they had in the past. Most (80%) of posts with the words *scared* and *tell* used the present tense, such as “I am scared to tell”, but 20% of posts used the past tense, such as saying “I was scared to tell”. The posts using the past tense tended to be sharing experiences for others, using phrases such as “i was scared… similarly like you”, but go on to share how they had had positive experiences and reassuring the original poster. From the survey, 29% of participants reported visiting the forum because they wanted to offer help, advice, or information to other forum users. Similarly, one theme from the interviews was that some users kept coming back to the forum to help others who were experiencing the same things they had experienced in the past. There was a sense of fulfilment from being able to help others, and a sense of giving back to the forum space. These findings show that there is a subgroup of forum users who continually return to the forum, sharing their experiences to help others. This complements previous research (47) which found that a small proportion (1%) of a domestic violence forum’s userbase were responsible for almost half of all posts. Having a group of people who continually return and post to the forum is essential to building an online community, even for people who only read the forum, rather than a collection of posts or isolated exchanges on a common topic (47).

This project focused on UK-based forums, including comparisons with UK national services, which may limit transferability to other settings. However, the structure of metainformation of the Magpie forums’ dataset which includes information on “user_country” suggests the forums’ considerable reach outside the UK, particularly in other English-speaking countries. Of the registered users who provided information on their country, 76% were UK-based and 20% were based in US, Australia, Canada and Ireland; however, there were users from an additional 105 countries. Even though the Magpie forums selected for this study were run by UK-based charities, the userbase included people from all over the world. Some countries may not have equivalent charities, so people seek support from other countries online instead. This reinforces the idea from previous research that forums overcome geographical isolation (19), but on a global scale. The interviews showed that some people searched for help online, and found open forums like Starling, Magpie or Chaffinch. In contrast, others were referred by therapists or school staff to online forums as a geographically commissioned service, such as Sparrow or Dunnock.

## Conclusion

Online forums for mental health are increasingly used across and beyond the UK. Although the survey indicated the forums were used by predominantly white and female users, there is some suggestion that forums attract more ethnic diversity, males, and younger people than NHS Talking Therapies. There is evidence that people who are questioning their gender identity or sexuality, neurodiverse people, and those with physical or geographical restrictions may feel more able to share their experiences on forums. This is because forums offer safe, anonymous spaces where people can share their fears, experience cathartic expression, connect with people with similar experiences, and give and receive support. These spaces are more accessible than health services, forming an important addition to offline support options.

##### Lived experience commentary by J.S. (forum moderator) and Neil Caton

Understanding who uses forums and why is crucial in providing insight into several key areas that can have wide-ranging impacts on forum users and the services directly or indirectly connected to them.

The mixed-methods approach has done well to capture and bring together some of the more intricate complexities involved. Some of these are not always easily recognised without the awareness of multiple perspectives, such as with the reasons behind preferences for online versus mainstream services and the ways in which these services may or may not interlink.

Moderators are in a unique position of being able to provide support much faster and to a broader range of people than many mainstream services. It is therefore encouraging to see highlighted, how forums and, by extension, their moderation have felt to be valuable, particularly for vulnerable and minority groups who may find mainstream services more inaccessible.

It is, however, important to consider that forums are being used predominantly by many of the same demographic who also use NHS Talking Therapies. This raises the question of what support is being used amongst people outside of this demographic, and whether there is a need to incorporate some of the benefits of forums and other alternative forms of support into mainstream services. This would allow forums to support a broader range of people and, if that is the case, to explore where it may be realistically possible to do so and how that could be achieved.

This research has particularly highlighted the significance of feeling connected within the support that is offered to people, but also that what constitutes feeling connected within the support is not a one-size-fits-all. This is a step in the right direction, which has the potential to lead to lasting changes that work for the current needs of a greater variety of people in a way that also provides more hope for the future, individually and collectively.

## Data Availability

The anonymised survey data is openly available from Lancaster University's repository PURE. The interview and forum post data are also stored on PURE, and available on request due to the potentially identifiable nature of the data.

https://doi.org/10.17635/lancaster/researchdata/714

